# Phenome-wide analysis of genetically imputed neuroimaging phenotypes reveals associations with psychiatric traits in a multi-ancestry cohort

**DOI:** 10.64898/2025.12.16.25342202

**Authors:** Lina Chihoub, Corinde E. Wiers, Joel Gelernter, Bingxin Zhao, Christal N. Davis, Henry R. Kranzler

## Abstract

**Background:** Understanding how variation in brain structure and function contributes to psychiatric and behavioral phenotypes remains a key challenge. The absence of neuroimaging data in many study samples limits this effort.

**Methods:** We used genome-wide association study (GWAS) summary statistics from the UK Biobank to impute 301 brain imaging-derived phenotype (IDP) genetic scores (IGS) in the Yale-Penn cohort, which is enriched for substance use disorders (n = 10,275; 52.8% European-like [EUR] and 47.2% African-like [AFR] genetic ancestry). The brain IDPs include white matter microstructure, regional volume, and resting-state functional connectivity measures, for which we generated IGS in the Yale-Penn participants. We then conducted a brain-wide phenome-wide association study (pheWAS) of the 301 IGS across 692 behavioral, psychiatric, and environmental traits.

**Results:** Among EUR individuals, we identified 19 IGS with significant associations that survived within-trait corrections for multiple testing. These included links between genetically predicted white matter integrity and sedative abuse, tobacco withdrawal, attention deficit hyperactivity disorder (ADHD); structural brain volumes and cocaine dependence, ADHD, and conduct disorder; and functional connectivity with substance-related symptoms and social phobia. Among AFR individuals, we identified 15 IDPs with significant associations, including associations between genetically predicted white matter integrity and stimulant use disorder, regional brain volumes and opioid withdrawal/dependence, and functional connectivity and cocaine craving.

**Conclusions:** Genetically imputed brain features capture biological variation associated with psychiatric traits. This work provides a framework for leveraging genetic data to link neuroimaging measures to substance use and mental health outcomes in samples that lack imaging data.

## Introduction

Structural and functional imaging techniques used to characterize the human brain have become increasingly powerful tools in psychiatric research. Imaging data have enabled the identification of biomarkers of psychiatric disorders by associating clinical traits with imaging-derived phenotypes (IDPs)—quantitative measurements reflecting white matter tract integrity, total and regional volumes, or functional network connectivity (1). For example, recent investigations of schizophrenia have identified cerebello-thalamo-cortical hyperconnectivity (1, 2) and reduced volume of the corpus callosum (3) as possible imaging biomarkers. Thus, the use of neuroimaging data may advance our understanding of etiological mechanisms and inform risk and treatment stratification of psychiatric disorders (3–5).

However, brain imaging techniques such as magnetic resonance imaging (MRI) are expensive and time consuming to perform at scale (6). Such challenges often limit the size of imaging cohorts, yielding insufficient statistical power to detect associations and reducing generalizability across populations (7–9). This has led to a reliance on datasets such as the UK Biobank (UKB), the world’s largest brain imaging study with over 100,000 participants. It features diffusion, structural, and resting-state functional MRI in addition to genotype data and electronic health records, making it a valuable resource for studying brain-behavior relationships (10, 11). However, the UKB is limited by the predominance of healthy volunteers of white British ancestry (12). Thus, it does not, on its own, support a comprehensive analysis of psychiatric traits across the diverse populations affected by these conditions.

One approach to address this limitation is integrating UKB imaging data with the phenotypic depth and diversity of non-imaged psychiatric cohorts. Recent efforts to bridge this gap have leveraged the heritable nature of brain IDPs—demonstrated through genome-wide association studies (GWAS) that link variation in these traits with common genetic variants (13–16)—to yield genetically derived proxy imaging measures. Notably, Yang et al. applied this approach to calculate IDP genetic scores (IGS)—an individual’s genetically estimated value of an IDP—in the All of Us Research Program and Alzheimer’s Disease Neuroimaging Initiative Study using GWAS summary statistics from the UKB (17). The resulting IGS were used to identify neurobiological differences associated with ocular, cardiovascular, and renal diseases, demonstrating the utility of genetically imputed brain IDPs for understanding disease risk (17).

We built on this work to estimate brain IGS and identify trait associations in the multi-ancestry, deeply phenotyped Yale-Penn cohort (N = 10,275), which is enriched for individuals with substance use disorders (SUDs) and co-occurring psychiatric disorders (18). We obtained effect estimates from UKB imaging GWAS to estimate IGS for 301 brain IDPs across three imaging domains (white matter microstructure, regional/total brain volumes, and functional network connectivity). We then conducted a brain-wide phenome-wide association study (pheWAS) across 692 psychiatric and behavioral traits in European-like (EUR) and African-like (AFR) genetic ancestry groups to uncover brain-behavior associations. This work expands the reach of imaging genetics in psychiatric research by leveraging genetic prediction and bypassing direct imaging measurement, thus enhancing the utility of datasets that lack neuroimaging.

## Methods and Materials

### Datasets

#### Brain Imaging Genetics Knowledge Portal (BIG-KP)

We obtained publicly available IGS weights derived from GWAS conducted in EUR individuals for 301 brain IDPs (https://zenodo.org/records/7709788). The IDPs spanned three imaging modalities (1) 110 white matter microstructure measurements (19); (2) 101 structural MRI (sMRI) values for total and regional brain volumes (20); and (3) 90 resting state mean functional MRI (rs-fMRI) activity values (21). These IDPs were chosen because they demonstrated significant out-of-sample prediction in previous research, with the amount of variance explained ranging from .0000024% (SE = 0.466%) to 4.72% (SE = 1.48%) for white matter microstructure, from 0.0762% (SE = 1.49%) to 7.01% (SE = 1.44%) for sMRI IDPs, and from .000015% (SE = 1.51%) to 5.02% (SE = 1.47%) for rfMRI traits (17). SNP-based heritability estimates for the white matter microstructure IDPs ranged from 22.4% (SE = 2.2%) to 66.5% (SE = 2.2%) (19), for sMRI IDPs from 5.6% (SE = 3.8%) to 72% (SE = 3.7%) (22), and for *rs-f*MRI IDPs from 3.6% (SE = 2.0%) to 46.1% (SE = 2.2%) (23).

#### Yale-Penn Sample

The Yale-Penn sample (N = 10,275; Table 1) was recruited and assessed at five U.S. academic study sites between 2000 and 2013, with Institutional Review Board approval at each site and informed consent obtained prior to the conduct of research procedures. Individuals with alcohol, cocaine, or opioid use disorders were enrolled through treatment services and local media advertisements. We also recruited control participants with no history of an SUD (18, 24). Participants completed the Semi-Structured Assessment for Drug Dependence and Alcoholism (SSADDA), a comprehensive interview that yields reliable SUD and mental health criteria and diagnoses (25, 26). The SSADDA asks participants regarding their substance use, demographics, and psychosocial/medical histories. We focused on 692 items from the SSADDA that were retained through clinician input and included psychiatric diagnoses, symptom-level information, treatments received, and length/number of disorder episodes (18).

**Table 1.** Sample characteristics (N = 10,275).

|  | **European Ancestry (n = 5424)** | **African Ancestry (n = 4851)** |
| --- | --- | --- |
|  | Mean (SD) or N (%) | Mean (SD) or N (%) |
| **Sex** |  |  |
| Female | 2326 (42.9%) | 2174 (44.8%) |
| Male | 3098 (57.1%) | 2677 (55.2%) |
| **Age** | 39.8 (12.9) | 41.5 (10.2) |
| **Race and ethnicity** |  |  |
| African American/Black, Hispanic | 53 (1%) | 166 (3.4%) |
| African American/Black, non-Hispanic | 58 (1.1%) | 4310 (88.8%) |
| White, Hispanic | 424 (7.8%) | 35 (0.7%) |
| White, non-Hispanic | 4550 (83.9%) | 21 (0.4%) |
| Other | 339 (6.3%) | 319 (6.6%) |
| **Marital status** |  |  |
| Divorced | 1061 (19.6%) | 768 (15.8%) |
| Married | 1134 (20.9%) | 702 (14.5%) |
| Never Married | 2900 (53.5%) | 2946 (60.7%) |
| Separated | 226 (4.2%) | 329 (6.8%) |
| Widowed | 103 (1.9%) | 106 (2.2%) |
| **Annual household income** |  |  |
| <\$9,999 | 2061 (38%) | 2380 (49.1%) |
| \$10,000 - 19,999 | 826 (15.2%) | 887 (18.3%) |
| \$20,000 - \$29,999 | 629 (11.6%) | 545 (11.2%) |
| \$30,000 - \$39,999 | 387 (7.1%) | 357 (7.4%) |
| \$40,000 - \$49,999 | 351 (6.5%) | 198 (4.1%) |
| \$50,000 - \$74,999 | 465 (8.6%) | 250 (5.2%) |
| \$75,000 - \$99,000 | 290 (5.3%) | 94 (1.9%) |
| \$100,000 + | 360 (6.7%) | 101 (2.1%) |
| **Education** |  |  |
| Less than high school | 1305 (24.1%) | 1627 (33.5%) |
| High school | 1228 (22.6%) | 1363 (28.1%) |
| Some college/technical school or higher | 2891 (53.3%) | 1860 (38.3%) |
| **Lifetime DSM-IV Substance dependence** |  |  |
| Alcohol | 2742 (50.6%) | 2493 (51.4%) |
| Cocaine | 2607 (48.1%) | 2907 (59.9%) |
| Marijuana | 1502 (27.7%) | 1318 (27.2%) |
| Opioids | 2216 (40.9%) | 917 (18.9%) |
| Sedatives | 542 (10%) | 129 (2.7%) |
| Stimulants | 362 (6.7%) | 140 (2.9%) |
| Tobacco | 2928 (54%) | 2507 (51.7%) |
| Other | 759 (14%) | 497 (10.2%) |
| **Mental health** |  |  |
| Antisocial personality disorder | 698 (12.9%) | 582 (12%) |
| Attention deficit hyperactivity disorder | 458 (8.4%) | 209 (4.3%) |
| Conduct disorder | 124 (2.3%) | 99 (2%) |
| Major depressive disorder | 837 (15.4%) | 529 (10.9%) |
| Manic episode | 33 (0.6%) | 25 (0.5%) |
| Panic disorder | 383 (7.1%) | 134 (2.8%) |
| Pathological gambling | 330 (6.1%) | 466 (9.6%) |
| Social phobia | 324 (6%) | 161 (3.3%) |
| Suicidal ideation | 2011 (37.1%) | 1517 (31.3%) |

*Note:* DSM-IV = Diagnostic and Statistical Manual, version 4. Percentages are based on available data; some categories do not sum to 100% due to missing or unknown values.

#### IGS Generation and PheWAS

Because the YalelZlPenn sample did not undergo brain imaging, we estimated IGS for the 301 brain IDPs using the --score command in PLINK (27, 28), which combines genotype data with GWAS effect size estimates to produce a single IGS per person and phenotype.

Posterior genetic variant effect sizes were trained using PRS-CS software (29). We evaluated the performance of the EUR-derived weights in the AFR group to facilitate inclusion of AFR individuals in the study despite the likelihood that applying variant weights based on the EUR GWAS to AFR individuals would reduce statistical power to detect IGS-phenotype links (30). The IGS were standardized within genetic ancestry groups.

We used the PheWAS package in R to conduct pheWAS across 692 traits for each IGS in the Yale-Penn cohort. Binary and continuous traits were regressed on each standardized IGS using logistic or linear regression models, respectively. Covariates included age, sex, and the first 10 within-ancestry genetic principal components computed using a principal component analysis (18). We controlled for multiple testing of the 692 traits in association with each IGS using the Benjamini-Hochberg false discovery rate (BH-FDR) procedure (q < 0.05), which given our interest in drawing IGS-specific inferences was our primary method for evaluating significance. We also applied more stringent corrections by pooling the 692 p-values per IDP across all IGS in each modality (i.e., 110 DTI, 101 sMRI, and 90 *rs-f*MRI).

#### Enrichment Analysis

To identify phenotypic domains that show more significant associations with a given modality than expected by chance, we performed a one-sided hypergeometric test within each genetic ancestry group and imaging modality. We categorized phenotypes by domains identified in previous work (18). Tests and significant associations (q < 0.05) were enumerated for each ancestry–modality stratum and phenotype domain. These counts, paired with the overall numbers of tests and significant results, were subsequently inputted to the hypergeometric function in R. Significance levels (corrected via the BH-FDR procedure), the number of significant associations expected by chance, and the enrichment ratio (i.e., the number of significant associations within each domain divided by the expected hits) were obtained.

## Results

### Sample Characteristics

Cohort characteristics are presented in **Table 1**. The Yale-Penn sample is balanced across EUR and AFR genetic ancestry groups and sex and is diverse in age and educational attainment. Household income is modest among most participants.

### European-like Ancestry Group

The pheWAS identified 19 IDPs of the 301 studied that were associated (q < 0.05) with at least one phenotype, for a total of 60 associations (Tables S1-S3; Figure 1). The associations spanned all three imaging modalities and a variety of phenotypic domains, particularly mood disorders, antisocial personality disorder (ASPD), attention deficit hyperactivity disorder (ADHD), conduct disorder, and SUDs. No associations survived the more stringent within-modality corrections for multiple testing.

**Figure 1:**
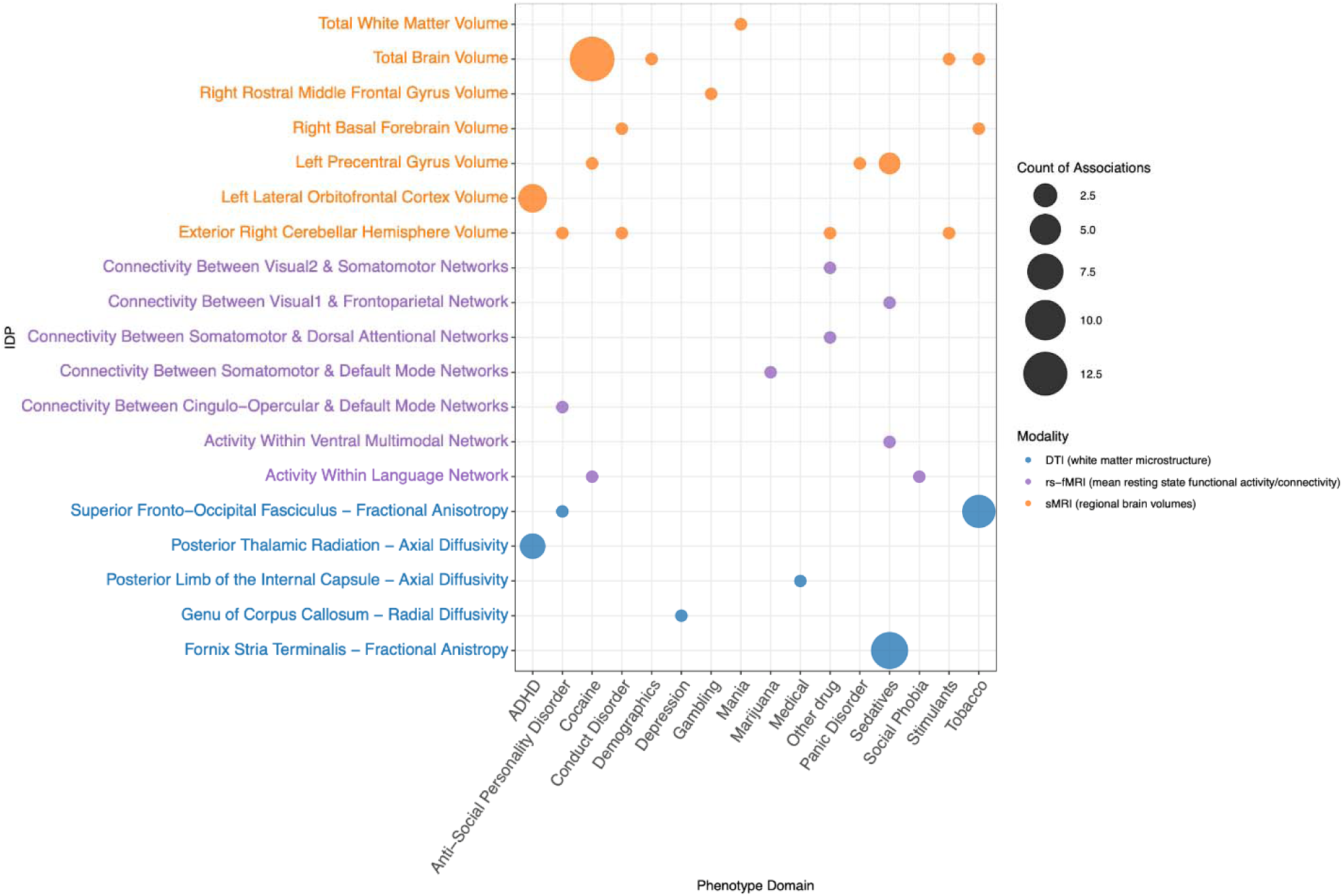
Summary of significant IGS-phenotype associations in European-like ancestry individuals from the Yale-Penn sample. The dot size reflects the number of associations between each imaging-derived phenotype genetic score (IGS) and phenotype domain. Dot and y-axis label color indicate the imaging modality of the IGS (diffusion tensor imaging [DTI], resting-state functional magnetic resonance imaging [rs-fMRI], or structural magnetic resonance imaging [sMRI]).

#### DTI

Five IDPs (radial diffusivity in the genu of the corpus callosum, axial diffusivity in the posterior limb of the internal capsule, axial diffusivity in the posterior thalamic radiation, fractional anisotropy [FA] in the superior fronto-occipital fasciculus, and FA in the fornix/stria terminalis) from the DTI modality survived within-IGS corrections for multiple testing, for 20 total associations. Links were observed between white-matter microstructure and depression, ADHD, stroke, and tobacco and sedative use. The tobacco-related associations typically involved negative links to withdrawal symptoms (e.g. irritability, anxiety), in which genetic liability for lower DTI measurements correlated with higher odds of withdrawal symptoms. For sedatives, higher FA in the fornix and stria terminalis corresponded to lower odds of meeting abuse criteria. Genetic liability for greater axial diffusivity in the posterior thalamic radiation was associated with a higher likelihood of exhibiting ADHD diagnostic criteria (e.g., duration of symptoms). The relationship between stroke and lower genetic liability to axial diffusivity in the posterior limb of the internal capsule was the most robust in this modality (β = −0.62, SE = 0.13, q = 0.001).

#### sMRI

Of the 101 regional and total brain volume traits tested, 7 were associated with phenotypes after within-IGS correction for multiple testing, corresponding to 32 associations. Most region-of-interest (ROI) IGS associations were with cocaine, with more limited links to sedative, tobacco, and stimulant use. Cocaine-related traits were consistently and strongly associated with IGS for reduced total brain volume. Additionally, experiencing two or more cocaine withdrawal symptoms was associated with lower IGS for volume of the left precentral gyrus. This IGS was also linked to sedative phenotypes. Another reward processing-related phenotype, pathological gambling, was associated with greater genetically predicted volume of the right rostral middle frontal gyrus.

ADHD, particularly impairment-related traits, was consistently associated with reduced genetically predicted volume of the left lateral orbitofrontal cortex. Conduct disorder symptoms (i.e., engaging in breaking and entering, truancy) were associated with lower IGS for volume of the right cerebellar hemisphere and greater IGS for volume of the right basal forebrain. ASPD criterion count was similarly associated with lower IGS for volume of the right cerebellar hemisphere, while bipolar I disorder was associated with higher IGS for total white matter volume.

#### rs-fMRI

Of the 90 mean functional connectivity IGS tested, associations with seven survived within-trait correction, yielding eight IGS-phenotype links. The connectivity IGS were mostly negatively associated with substance-related traits (e.g., cocaine, marijuana, sedatives) across various brain networks (e.g. sensorimotor control, attentional control, visuomotor integration). Sedative dependence was linked to lower IGS for functional connectivity between the primary visual and frontoparietal networks, which was the most robust association among this modality group in the EUR cohort. Although most associations were negative, suggesting that genetic liability for lower functional connectivity between or within the networks was correlated with higher odds of the phenotype, there were exceptions. These included the association between lower IGS for connectivity within the language network and an earlier age of social phobia diagnosis. Additionally, lower IGS for connectivity between the cingulo-opercular and default-mode network was associated with a lower likelihood of being arrested in participants with ASPD.

### African-like Ancestry Group

In the AFR cohort, 15 IGS were associated with at least one phenotype, for a total of 97 associations (Tables S4-S6; Figure 2). Most associations were with substance-related traits, particularly opioids and stimulants. Less prominent were associations with mental health traits, including ASPD and suicidal behaviors, and neurodevelopmental disorders. As in the EUR cohort, no associations survived the more stringent within-modality corrections.

**Figure 2:**
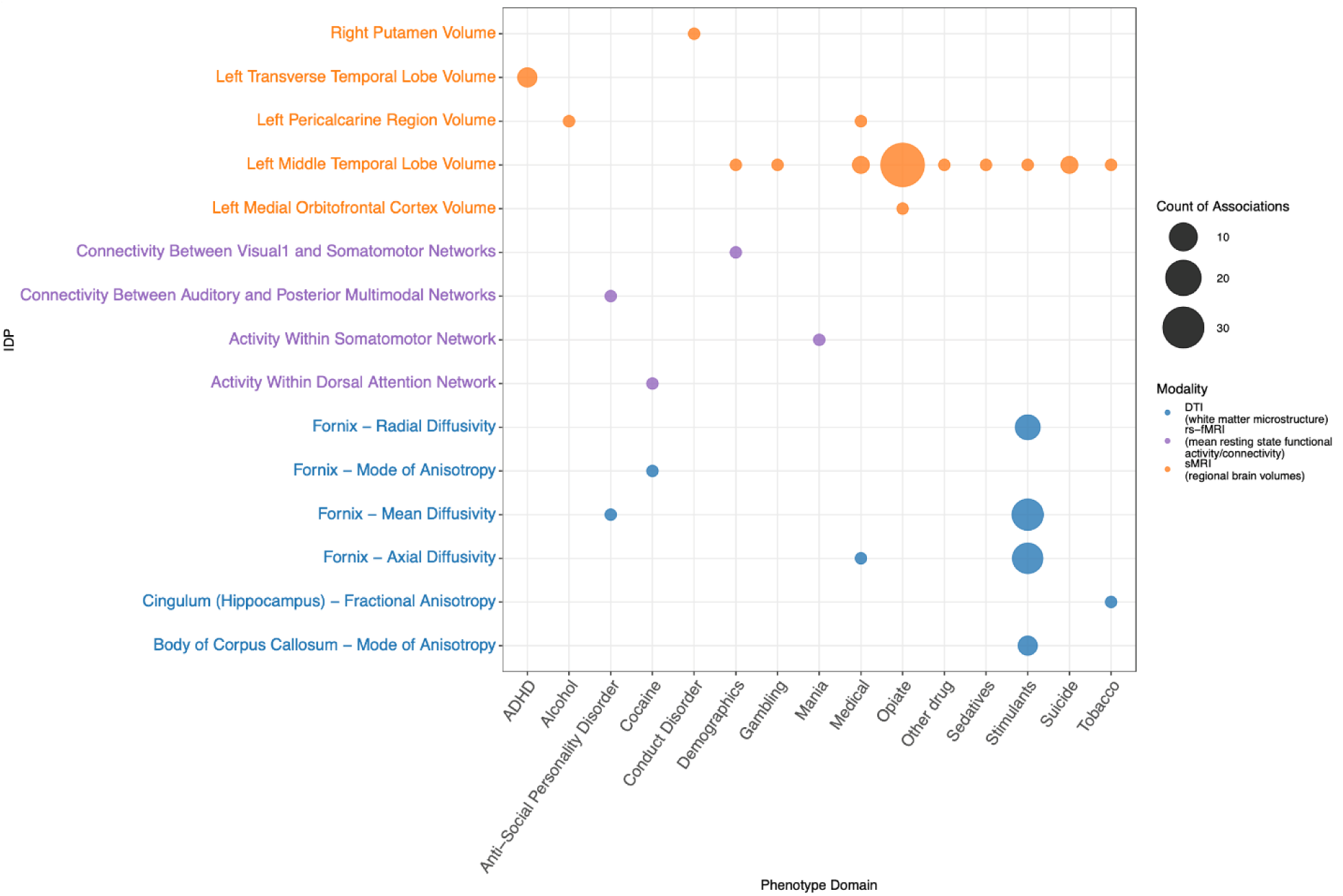
Summary of significant IGS-phenotype associations in African-like ancestry individuals from the Yale-Penn sample. The dot size reflects the number of associations between each imaging-derived phenotype genetic score (IGS) and phenotype domain. Dot and y-axis label color indicate the imaging modality of the IGS (diffusion tensor imaging [DTI], resting-state functional magnetic resonance imaging [rs-fMRI], or structural magnetic resonance imaging [sMRI]).

#### DTI

Six IGS had links that survived within-IGS correction, for a total of 41 significant trait associations. Stimulant-related traits dominated this category. A higher IGS for mode of anisotropy in the corpus callosum was associated with lower propensity for stimulant use disorder and dependence (β = −0.28, SE = 0.07, q = 0.02 and β = −0.33, SE = 0.08, q = 0.02, respectively). Greater genetically predicted axial and radial diffusivity in the fornix were associated with stimulant use, abuse, and dependence. The link between axial diffusivity in the fornix and stimulant abuse constituted the strongest relationship of the AFR DTI group (β = 0.36, SE = 0.08, q = 0.002). Greater genetically predicted mean diffusivity in the fornix was similarly positively, robustly associated with stimulant-related traits. Mode of anisotropy in the fornix IGS was linked to a later age of first cocaine use and the IGS for greater mean diffusivity in the fornix was linked to acting out behaviors in ASPD.

#### sMRI

Five IGS were significantly associated with at least one phenotype, for a total of 52 associations. Lower genetically predicted volume of the left middle temporal lobe was strongly linked to a variety of opioid-related traits, including the number of times used; the frequency of use; the mental, social, and cognitive consequences of use; withdrawal symptoms; dependence diagnosis; and abuse criteria. The IGS for this region was also linked to tobacco and sedative dependence and stimulant withdrawal. There were also positive associations between the left middle temporal lobe IGS and lifetime suicide attempt and suicidal intent.

The left transverse temporal lobe IGS had the most robust associations within the AFR group. Lower genetically predicted volume in this region was associated with greater odds of endorsing ADHD-related impairment in multiple settings and prolonged ADHD symptom duration. There were also an inverse association between genetically predicted volume of the left pericalcarine region and years of opioid use, and a positive association between volume of the right putamen and acting-out behaviors in conduct disorder.

#### rs-fMRI

Four rs-fMRI IGS survived within-IGS corrections, for a total of four associations. The most robust association was between greater genetically predicted mean functional connectivity within the dorsal attention network and cocaine craving. Additionally, lower IGS for functional connectivity within the somatomotor network was linked to a longer duration of mania episodes. Greater genetically predicted connectivity between the auditory and posterior multimodal networks was linked to impulsive behavior in ASPD.

### Enrichment Testing

#### European-like Ancestry

Five phenotype domains were enriched (q < 0.05) for associations with an IGS. Sedative (enrichment = 6.3, p = 0.0004), tobacco (enrichment = 6.93, p = 0.002), and ADHD (enrichment = 9.45, p = 0.03) phenotypes showed enriched associations with DTI IGS. For the ROI IGS, cocaine phenotypes showed enrichment (enrichment ratio = 7.0, p = 5.28 lZl 10^-8^) as did ADHD (enrichment ratio = 7.86, p = 0.02).

#### African-like Ancestry

Two phenotype domains were enriched in the AFR genetic ancestry group: stimulant phenotypes in association with DTI IGS (enrichment ratio = 16.0, p = 1.11 lZl 10^-40^), and opioid phenotypes with ROI IGS (enrichment ratio = 9.2, p = 2.46 lZl 10^-27^).

## Discussion

We leveraged genetic imputation of IGS for 301 neuroimaging traits spanning white matter microstructure, regional and total brain volumes, and resting-state functional connectivity to examine potential neural substrates of psychiatric and substance use traits in the Yale-Penn cohort. Despite deep phenotyping for psychiatric traits in this cohort, it lacked imaging data, so that imputed measures derived from large-scale imaging GWAS provided informative proxies. We identified 157 significant IGS-phenotype associations in EUR and AFR individuals. Across ancestry groups and imaging modalities, these associations most frequently involved substance use and externalizing traits. Although this likely reflects the characteristics of the Yale-Penn cohort, which was ascertained for substance use, it also provides important insights into neural signatures of disinhibited behaviors. Together, these findings demonstrate that genetically imputed IDPs capture meaningful variation in neural structure and function relevant to substance use, externalizing behaviors, and other psychiatric traits, and that these associations can be identified in the absence of direct neuroimaging data.

### DTI Modality

Among AFR individuals, greater IGS for mean, axial, and radial diffusivity in the fornix were associated almost exclusively with stimulant traits, while in EUR individuals, there were links between FA measures in the fornix and sedative abuse. Despite phenotypic divergence, findings in both ancestry groups consistently show that genetically predicted variations in the fornix were associated with substance-related traits. These findings parallel neuroimaging studies that have generally shown alterations in diffusivity across major white matter tracts in substance-using individuals (31), particularly those who use methamphetamine (32–34). The fornix is classically associated with memory formation and may shape reward-seeking behavior (35). Its consistent associations across substances and ancestry groups suggest that alterations in white matter integrity in this tract may be a common underlying neural substrate among individuals who use substances and those with SUDs. Notably, previous neuroimaging studies of the effects of substance use have generally reported *reduced* FA in the fornix (31, 36–38), whereas we found *greater* genetically predicted FA values with sedative abuse. However, the direction of FA change appears to depend on the type of substance used, the age of exposure, the duration and severity of use, and presence of comorbid psychiatric conditions (39, 40).

### sMRI Modality

In AFR individuals, lower IGS for gray matter volume in the left middle temporal lobe was associated with several opioid use and withdrawal symptoms. Lower gray matter volume in the temporal lobe has been demonstrated in individuals who use opioids (41), and our findings add genetic support to the potential involvement of this specific subregion in opioid use (42, 43). The middle temporal lobe is classically linked to semantic cognition, sensory integration, and language processing (44), but its role in opioid use may relate to attentional and cue-based biases that promote craving, possibly mediated by the default mode network (42, 45, 46). Notably, we also found that the same region was negatively associated with stimulant withdrawal symptoms, duration of sedative use, and tobacco dependence. Thus, the left middle temporal gyrus may represent a convergent site of vulnerability across multiple SUDs (47–51).

In EUR individuals, genetically predicted lower total brain volume was associated with cocaine use, which aligns with prior evidence of gray and white matter loss in cocaine dependence (52–60). Genetically predicted lower total brain volume was also associated with tobacco use and stimulant dependence. As these findings are consistent with total gray and white matter volumetric loss reported in prior imaging studies (61–64), they may reflect a structural signature for substances with stimulating effects.

Beyond substance use, several ROI associations pointed to structural correlates of externalizing and developmental traits. In EUR individuals, the right rostral middle frontal gyrus IGS was linked to gambling disorder, while across EUR and AFR individuals IGS for the right basal forebrain, exterior right cerebellar hemisphere, and right putamen were associated with conduct disorder. Several of these regions are involved in reward valuation and impulsivity (65–72), implicating fronto-striatal structures in disinhibitory and antisocial behaviors.

In AFR individuals, genetically predicted lower volume in the left transverse temporal lobe (Heschl’s gyrus) was associated with ADHD criteria, consistent with previous studies (73–76). Whereas Heschl’s gyrus is involved in auditory processing (77), this finding aligns with evidence linking auditory processing deficits to attentional dysregulation in ADHD (78) and volumetric differences in Heschl’s gyrus may serve as a biomarker for the disorder (73). In contrast, among EUR individuals, associations with ADHD were localized to the left lateral orbitofrontal cortex, a region with roles in goal-directed and motivational behaviors (79) that has been implicated in structural imaging studies of ADHD (80, 81). These findings highlight neural substrates associated with attentional and behavioral regulation, reinforcing evidence of involvement of the temporal and orbitofrontal cortical regions.

### *rs-f*MRI Modality

There were fewer associations with the *rs-f*MRI IGS than with the DTI and sMRI IGS. In AFR individuals, greater genetically predicted connectivity within the dorsal attention network was associated with cocaine craving, consistent with a potential conceptualization of craving as a shift in goal-directed attention toward drug-related cues (82, 83). This observation also aligns with an imaging study of formerly cocaine-dependent individuals, which showed reduced resting-state within-network connectivity in the dorsal attention network (84).

In EUR individuals, genetically predicted lower connectivity within the language network was associated with an earlier onset of social phobia. Although social phobia has most commonly been associated with the emotional and attentional networks (85–87), some studies have reported that individuals with the disorder show reduced resting-state functional connectivity in regions implicated in speech initiation, the interpretation of social and linguistic information, and self-referential processing (88–91). Thus, a genetic predisposition to impaired integration of sociolinguistic information may contribute to an earlier development of social phobia.

Greater genetically predicted connectivity between the cingulo-opercular and default mode networks was linked to antisocial behaviors in EUR individuals. Although connectivity between these networks has not yet been implicated in ASPD, there is evidence supporting their individual roles (92–94). Engagement of the cingulo-opercular network during goal-directed or executive processes may normally be accompanied by reduced self-referential activity in the default mode network (95). Thus, greater connectivity between these networks suggests that a deficit in network switching ability could contribute to impulsive behaviors. Thus, collectively, genetically imputed connectivity patterns may index meaningful variation relevant to both internalizing (e.g., social phobia) and externalizing behaviors (e.g., ASPD).

### Conceptual and Methodological Implications

Genetically imputed imaging traits show potential to identify biologically coherent, transdiagnostic patterns and can complement traditional imaging studies. Whereas the conventional approach of directly imaging cohorts is limited by cost, time, and accessibility (6), using genetics to impute these traits constitutes a rapid and scalable alternative that can increase cohort diversity and sample size. Thus, the approach helps to address two long-standing limitations of brain-wide association studies (7, 8). First, the continued overrepresentation of White participants and those from higher socioeconomic and educational backgrounds confounds much psychiatric neuroimaging work, despite evidence that these factors may moderate associations between clinical traits and imaging features (9, 96).

Additionally, efforts to identify imaging biomarkers for psychiatric traits have been limited by the nonspecific, overlapping nature of many reported findings (3, 97). By estimating IGS in deeply phenotyped cohorts, it may be possible to enhance biomarker specificity. This would better align with emerging conceptualizations of psychiatric research, such as the Research Domain Criteria, which emphasize dimensional, functional domains over categorical diagnoses (98, 99).

The use of genetically imputed IDPs also can inform our understanding of the genetic contributions to neural correlates of substance use and mental health disorders. However, the precise etiology of the identified IGS-trait relationships requires further research. One possibility is that the same genetic factors may independently influence brain structure/function and psychiatric traits. Alternatively, genetic variants may influence neuroanatomical features, and in turn increase risk for psychiatric conditions. Studies that incorporate Mendelian randomization could evaluate potential directional pathways linking brain phenotypes and psychiatric traits (100). Additionally, genetic correlation and other analyses of pleiotropy could help to quantify the extent of shared genetic architectures between brain structure/function and psychiatric traits (101, 102). Resolving the mechanisms that underlie the identified relationships may help to inform more precise, biologically based frameworks for understanding and treating psychiatric conditions.

Finally, comparing genetically predicted imaging traits with directly measured imaging traits in samples with both can yield more precise estimates of environmental influences, such as substance use, on the brain. This approach may facilitate a greater understanding of the potential neurotoxic effects of abused substances than would otherwise be possible.

## Limitations

In addition to its strengths, our study had several limitations, including the unavailability of GWAS for IDPs in AFR individuals. Differences in allele frequencies and linkage disequilibrium structure between ancestry groups limit the utility of the EUR IGS weights as proxies of brain structure and function in AFR individuals, which reduces the power to detect IGS-phenotype associations among AFR individuals (30). Additionally, IGS are not equivalent to observed brain imaging measures and account for only a proportion of the variance in such measures (17). Thus, findings must be interpreted as reflecting genetic liability. Finally, no IGS-phenotype associations were significant after correcting for within-modality multiple testing. This reflects both the modest effect sizes and large number of tests performed. As larger GWAS of IDPs become available, the predictive utility of IGS should increase.

## Conclusions

By integrating genetic imputation of IDPs with deep phenotyping, we identified 157 associations predominantly with substance use and externalizing traits. Results converged on the relevance of behavioral regulation, attentional, and sensorimotor neural systems across ancestries and imaging modalities. This work offers a practical approach to address the longstanding question of how variation in brain structure and function underlies neuropsychiatric conditions. As our understanding of the links between genetic variation, neural phenotypes, and behavior evolves, this approach may facilitate more precise, biologically informed models of psychiatric vulnerability.

## Data Availability

IGS weights used in the study are publicly available at https://zenodo.org/records/7709788.

## Acknowledgments

This research was supported by the Veterans Integrated Service Network 4 Mental Illness Research, Education and Clinical Center and by grants I01 BX004820 and R01 AA030056 to HRK.

